# Predictors of Treatment Response, Remission, Relapse and Rehospitalization in First-Episode Psychosis Patients in Medellin, Colombia

**DOI:** 10.1101/2021.11.22.21266701

**Authors:** Jenny Garcia, Lina M. Agudelo, Maria A. Canas, Natalia Castro-Campos, Oscar J. Ribero, Juan A. Gallego

## Abstract

**Background:** Most studies with first episode psychosis patients have been conducted in high-income countries. On the other hand, very few first episode studies have been conducted in Latin-America. Therefore, the goal of our study is to determine predictors of treatment response, remission, relapse and rehospitalization in a first episode psychosis population from Medellin, Colombia.

**Methods:** Data was obtained from electronic health records from first episode patients with a diagnosis of schizophrenia-spectrum disorder diagnoses who were evaluated between January 2014 and December 2016 at two psychiatric institutions in Medellin, Colombia. Survival and Cox proportional hazard models were used for the analysis.

**Results:** Duration of untreated psychosis and hospitalization at the time of presentation was associated with both response and remission. Response was also predicted by less first-order symptoms and more years of education. Remission was predicted by older age of onset. Relapse and rehospitalization were predicted by use of substances and poor adherence to pharmacologic treatment. Less years of education and hospitalization at the time of presentation were also associated with rehospitalization.

**Conclusion:** Predictors of treatment response, remission, relapse and rehospitalization in first-episode patients are similar in Colombia compared to other high-income regions such as the United States and Europe.

## 1. Introduction

Schizophrenia is an illness with a prevalence rate of 0.33 to 0.75% of the population (Saha et al., 2005; Moreno-Küstner et al., 2018). Despite the relatively low prevalence rates, it is considered the 15th cause of years lived with disability in the world (GBD., 2016) and is associated with premature mortality (Olfson et al., 2015) and higher financial burden (Desai et al., 2013). For the past several years, the field has been conducting studies with patients who present with a first episode (FE) of the illness to study neuropathophysiological abnormalities, to determine the efficacy and safety of treatments in the absence of confounding factors such as cumulative effect of antipsychotic treatment or a long duration of illness and to investigate the impact of early interventions in the long-term outcome of schizophrenia.

Most studies with first episode schizophrenia patients have been conducted in the US (Schooler et al., 2005; McEvoy et al., 2007; Robinson et al., 2015; Kane et al., 2015), Europe (Kahn et al., 2008; Crespo-Facorro et al., 2013) and South Africa (Emsley et al., 1999). Accordingly, the evidence regarding predictors of either treatment response, remission, relapse or rehospitalization are derived from those studies. For example, a shorter duration of untreated psychosis (DUP) has been consistently found as a predictor of good response to treatment (Perkins et al., 2004) and/or remission of symptoms (Lieberman et al., 1996; Robinson et al., 2004; Emsley et al., 2007). Similarly, good premorbid functioning has been associated with good clinical response (Perkins et al., 2004; Rabinowitz et al., 2006; Crespo-Facorro et al., 2007). On the contrary, an early age of onset, poor adherence to treatment and comorbid substance use disorders have been associated with worse clinical outcomes, including relapse and rehospitalization (Robinson et al., 1999; Crespo-Facorro et al., 2007).

Unfortunately, even though Latin America comprises 20 countries and more than 600 million people, very few longitudinal FE studies have been conducted and only a handful of specialized FE teams have been established, mostly in Brazil, Chile and Mexico (Aceituno et al., 2020). Based on our literature review, we just found three clinical trials (Apiquian et al., 2003; Valencia et al., 2012; Valencia et al., 2017), few reports describing a naturalistic follow-up of clinical samples (Attux et al., 2007; Trzesniak et al., 2012; Gonzalez-Valderrama., 2017; Cano et al., 2021), a number of short, acute treatment studies used to examine neuroimaging or genetic markers (De La Fuente-Sandoval et al., 2013; Santoro et al, 2018) or chart reviews with a longitudinal component (Gomez-de-Regil et al., 2010; Markkula et al., 2011; Mena et al., 2018).

Given the small number of studies, only few demographic and illness predictors of clinical outcomes have been reported. Gomez-de-Regil and colleagues (2010) found that residual symptoms of psychosis were associated with a lower level of education and an insidious onset of symptoms. Mena and colleagues (2018) found that males with a younger onset of psychosis were over-represented in a group of first episode schizophrenia (FES) patients who received clozapine. Cano and colleagues (2021) followed 50 FES patients naturalistically for up to 5 years and found that poor functioning was predicted by a higher number of relapses, hospitalizations, and changes in the antipsychotic regimen. Lastly, Gonzalez-Valderrama and colleagues (2017) found that a shorter DUP was associated with a more robust improvement in negative symptoms.

Therefore, to expand on the current body of work, we present longitudinal data obtained from electronic health records (EHR) from a large FEP sample from Medellin, Colombia, to determine sociodemographic, illness and treatment predictive factors associated with treatment response, remission of symptoms, relapse and rehospitalization. Given that factors such as trauma, violence, and poverty have been associated with schizophrenia, and that these factors are commonly present in Latin-American countries, we hypothesize that other predictive factors may be more prominent in Latin American FE samples compared to other higher income countries.

## 2. Methods

### 2.1 Data extraction

Data was extracted EHR from two institutions in Medellin, Colombia: 1) the Hospital Mental de Antioquia, and 2) Clinica Samein. Both institutions serve patients from all socioeconomic classes and have a psychiatric emergency room, inpatient and outpatient services. Inclusion criteria: 1) Ages 65 or younger; 2) presented for psychiatric treatment for the first time between January 1^st^ 2014 and December 31^st^ 2016; 3) ICD-10 diagnosis of schizophrenia (F20), schizoaffective disorder (F25) or brief psychotic disorder (F23). The following exclusion criteria were applied: 1) Dementia; 2) Moderate and severe intellectual disability; 3) ICD-10 diagnosis of substance-induced psychosis (F19); 4) psychosis due to a general medical condition.

For each patient, the following information was extracted directly from the EHR based on the initial and subsequent psychiatric evaluations and for a period of up to 12 months in each individual: socio-demographic information, psychiatric diagnosis, psychiatric symptoms, treatment received and presence or absence of hospitalizations. Information was extracted, reviewed and entered in a database by J.G and L.A. Of note, there was no direct contact with patients. All information was obtained from the EHR. This protocol was reviewed and approved by the ethics committee at both institutions.

### 2.2 Outcome Definition

Response, remission, relapse and rehospitalization were the outcomes of interest in the study. To determine the outcomes, all psychiatric notes for each patient for a period of up to 12 months were reviewed by J.G, O.R. and L.A. Based on this information, the following definition was coded for each outcome: 1) response was coded when the treating psychiatrist reported that patient’s symptoms were improving; 2) remission was coded when the treating psychiatrist reported that positive symptoms of psychosis were no longer present 3) relapse was defined as a return of psychotic symptoms in those patients who had initially remitted according to the notes from the treating psychiatrist 4) rehospitalization was coded when patients had a second hospitalization after an initial hospitalization or a first hospitalization if they had not been initially hospitalized.

### 2.3 Statistical Analysis

Patients were grouped based on diagnostic categories (schizophrenia, schizoaffective disorder and brief psychotic disorder). Continuous variables were summarized using mean and SD when normally distributed or median and interquartile range if not normally distributed. An Anova test was used to compare continuous variables across the three groups. Categorical variables were compared using contingency tables and chi-square tests. Lastly, a multivariate Cox hazard regression model was calculated for each of the four outcomes. All relevant clinical variables with potential effects on the outcome were entered in the model.

## 3. Results

### 3.1 Subjects included

One-thousand-five-hundred and ninety-three patients (1593) patients who were evaluated for the first time between January 1^st^ 2014 and December 31^st^ 2016 were first identified. Of those, 805 (62.7%) were excluded for having a prior psychiatric visit at other institutions, 363 (28.3%) for not having the selected ICD-10 codes, 76 (5.9%) for having intellectual disability, 18 for having dementia (1.4%), 16 (1.2%) for having delusional disorder, and 5 (0.4%) for having schizotypal personality disorder. Three hundred and ten patients were finally included in the analysis (Figure 1).

**Figure 1.**
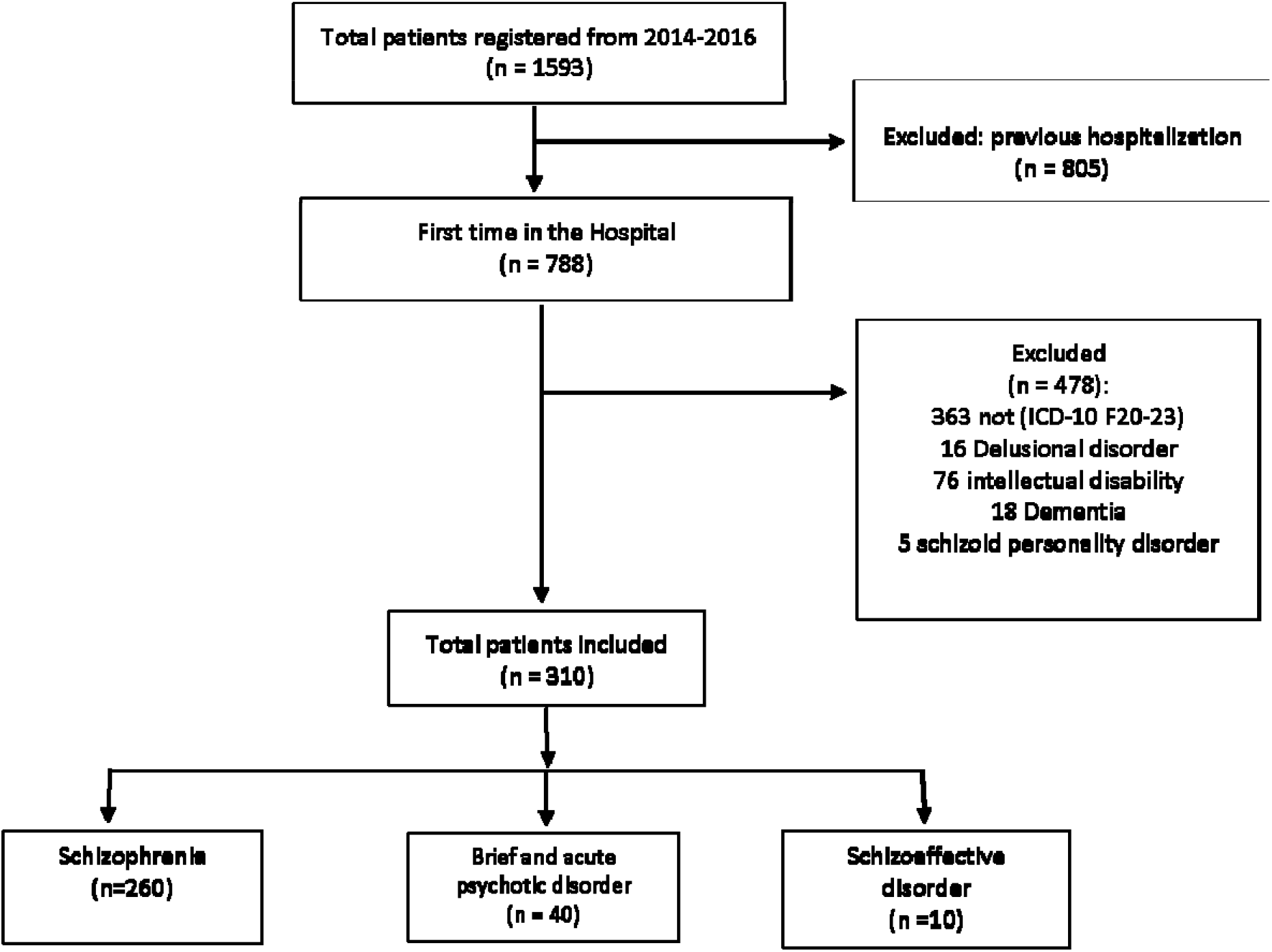
Flow diagram of patients entered into the study

### 3.2 Subject Characteristics

In the overall group, patients were mostly male 73.5%, single (80.1%) and had a median age of 25 years (IQR=19-38). They had been in school for a median of 11 years (IQR=7-11) and had a median duration of psychotic symptoms of 120 days (IQR=25.5-720). Forty one percent had low socio-economic status, 90.3% were living with family and 58.6% were hospitalized at the time of first presentation. Of the 310 patients included, 260 (83.9%) had schizophrenia, 10 (3.2%) had schizoaffective disorder and 40 (12.9%) had a brief psychotic disorder. The three groups were largely similar except for patients in the brief psychotic disorder group who had, compared to patients in the SZ and SAD groups, significantly shorter duration of psychotic symptoms (p=0.0001), lower frequency of first-order symptoms (p=0.01), and less blunted/flat affect (<0.001), alogia (p=0.037) and abulia (p=0.13) (Table 1).

**Table 1.** Baseline Characteristics by Diagnostic Groups

|  | Overall<br>(n=310) | Schizophrenia<br>(n=260) | Schizoaffective disorder<br>(n=10) | Brief Psychotic Disorder<br>(n=40) | p-value |
| --- | --- | --- | --- | --- | --- |
|  | n (%) | n (%) | n (%) | n (%) |  |
| Male, n(%) | 228(73.5) | 194(74.6) | 8 (80.0) | 26(65.0) | 0.39 |
| Age in years, median (IQR) | 25(19-38) | 25(19-37) | 35.5(24-48) | 25(19.5-38) | 0.34 |
| Single, n(%) [n=302] | 242 (80.1) | 207 (71.8) | 7 (77.8) | 28 (77.5) | 0.36 |
| Years of Education, median (IQR) | 11(7-11) | 11(7-11) | 9(7-11) | 11(7-13) | 0.76 |
| Duration of untreated psychosis in days, median (IQR) | 120(25.5-720) | 180(60-720) | 180(90-300) | 14(7-15) | <0.001 |
| Middle/high vs. Low SES, n(%) | 180(58.4) | 156(60.2) | 5(50) | 19(48.7) | 0.34 |
| Urban vs. rural residence, n(%) | 289 (93.2) | 243 (93.5) | 8 (80) | 38 (95) |  |
| Living situation, n(%) |  |  |  |  | 0.66 |
| Alone | 12 (3.9) | 10 (3.9) | 0 (0) | 2 (5) |  |
| With family | 280 (90.3) | 236 (90.8) | 10 (100) | 31 (85) |  |
| Institutionalized | 10 (3.2) | 7 (2.7) | 0 (0) | 3 (7.5) |  |
| Homeless | 3 (1.0) | 3 (1.2) | 0 (0) | 0 (0) |  |
| Hospitalized at presentation [309], n(%) | 181 (58.6) | 155 (59.6) | 8 (80) | 18 (45) | 0.11 |
| Hallucinations, n(%) | 287(92.6) | 240(92.3) | 9(90.0) | 38(95.0) | 0.79 |
| Delusions, n(%) | 301 (97.1) | 251 (96.5) | 10 (100) | 40 (100) | 0.41 |
| First-order symptoms, n(%) [309] | 197 (63.8) | 174 (67.2) | 6 (60) | 17 (42.5) | 0.01 |
| Disorganized thinking, n(%) [309] | 193 (62.5) | 164 (63.1) | 7 (70) | 22 (56.4) | 0.64 |
| Disorganized behavior, n(%) [309] | 184 (59.6) | 154 (59.5) | 6 (60) | 24 (60) | 0.99 |
| Catatonic symptoms, n(%) | 16 (5.2) | 14 (5.4) | 1 (10) | 1 (2.5) | 0.58 |
| Negative Symptoms |  |  |  |  |  |
| Blunted or flat affect, n(%) | 192 (61.9) | 173 (66.5) | 5 (50) | 14 (35) | <0.001 |
| Alogia, n(%) | 81 (26.1) | 75 (28.9) | 2 (20) | 4(10) | 0.037 |
| Abulia, n(%) | 111 (35.8) | 101 (38.8) | 4 (40) | 6 (15) | 0.013 |
| Aggressive behavior, n(%) | 186 (60) | 157 (60.4) | 8 (80) | 21 (52.5) | 0.27 |
| Insomnia, n(%) | 206 (66.5) | 172 (66.2) | 7 (70) | 27 (67.5) | 0.96 |
| Substance use disorder |  |  |  |  |  |
| Alcohol use disorder, n(%) [308] | 20 (6.5) | 15 (5.8) | 2 (20) | 3 (7.5) | 0.20 |
| Cannabis use disorder, n(%) [308] | 127 (41.2) | 113 (43.8) | 4 (40) | 10 (25) | 0.08 |
| Cocaine use disorder, n(%) [307] | 81 (26.4) | 71 (27.6) | 2 (20) | 8 (20) | 0.53 |
| Hallucinogen use disorder, n(%) [308] | 63 (20.5) | 56 (21x.7) | 0 (0) | 7 (17.5) | 0.22 |
| Inhalants, n(%) [306] | 20(6.5) | 19(7.4) | 0(0) | 1(2.6) | 0.37 |

SES: Socioeconomic status

### 3.3 Treatment characteristics

The most commonly used antipsychotic medication was olanzapine (60.2%), followed by risperidone (46.3%) and haloperidol (13.6%). Interestingly, a good number of patients were treated with risperidone long-acting injectable (LAI) (11.4%), pipotiazine palmitate LAI (7,1%) and clozapine (6.8%). In addition, about a quarter of patients were treated with two or more antipsychotic medications (23.3%), antidepressants (23.9%) or mood stabilizers (25.6%). 37.2% of patients were treated with benzodiazepines (Table 2).

**Table 2.** Treatment Characteristics of the Overall Sample

| Antipsychotic | N(%) | Median dose(IQR) | Mean dose (SD) |
| --- | --- | --- | --- |
| Olanzapine | 186(60.2) | 10(10-20) | 12.6(5.7) |
| Risperidone | 143(46.3) | 3(2-4) | 3.0(1.6) |
| Haloperidol | 42(13.6) | 5(5-10) | 6.7(4.9) |
| Risperidone (LAI) | 35(11.4) | 37.5(25-50) | 37.1(13.7) |
| Pipotiazine Palmitate (LAI) | 22(7.1) | 25(25-25) | 26.1(5.3) |
| Clozapine | 21(6.8) | 200(100-300) | 215.5(135.0) |
| Quetiapine | 16(5.2) | 100(25-150) | 128.1(146.9) |
| Paliperidone Palmitate (LAI) | 4(1.3) | 112.5(75-150) | 112.5(443.3) |
| Aripiprazole | 3(0.97) | 30(15-30) | 25.0(8.7) |
| Amisulpride | 2(0.6) | 300(200-400) | 300(141.4) |
| Paliperidone | 1(0.3) | 6(6-6) | 6(NA) |
| Asenapine | 1(0.3) | 10(10-10) | 10(NA) |
| APP | 72(23.3) | NA | NA |
| Antidepressants | 73(23.9) | NA | NA |
| Mood Stabilizers | 79(25.6) | NA | NA |
| Benzodiazepines | 115(37.2) | NA | NA |
| Antiparkinsonian medications | 52(16.8) | NA | NA |

LAI: Long Acting Injectable; APP: Antipsychotic Polypharmacy

### 3.4 Predictors

As described previously and observed in table 1, patients with brief psychotic disorder differed from the schizophrenia and schizoaffective disorder groups in several baseline variables. Moreover, out of the 40 patients with a brief psychotic disorder diagnosis, 23 dropped out of treatment either right after the baseline visit or right after hospitalization; therefore, only 17 of those patients provided follow-up data. Furthermore, all 17 patients achieved full remission and were not re-hospitalized. Therefore, we decided to exclude this group from the regression analysis and restrict our analysis to patients with schizophrenia and schizoaffective disorder. Lastly, we further excluded 53 patients from the regression analysis, given that they only provided data from the baseline visit.

#### 3.4.1 Treatment response

two hundred and one out of 232 patients responded to treatment (86.6%). Higher response rates were associated with duration of psychosis ≤6 months (adjHR=0.58, 95%CI=0.40-0.78, p<0.0001), lower frequency of first-order symptoms (adjHR=0.52, 95%CI: 0.34-0.81, p=0.004, hospitalization at the time of presentation (adjHR=3.69, 95%CI: 2.29-5.91, p<0.0001) and ≥ eleven years of education (adjHR=1.51, 1.01-2.24, p=0.04) (Supplemental table 1)

#### 3.4.2 Remission

One hundred thirty five of 260 (58.2%) patients achieved remission. Median time to remission of symptoms was 32 days (IQR: 17-118). As observed with treatment response, a duration of psychosis ≤ 6 months was associated with increased rates of remission (adjHR=0.52, 95% CI: 0.34 – 0.79, p=0.002) along with an older age of onset (adjHR=0.47, 95% CI: 0.28 – 0.82, p=0.007) and hospitalization at the time of presentation (adjHR: 2.32, 95% CI:1.39-3.88, p=0.001) (Supplemental table 2)

#### 3.4.3 Relapse

Out of the 201 patients who responded to treatment, 88 (43.8%) relapsed. Median time to relapse was 218 days (IQR: 90-369) with a median number of relapses of 1 (IQR: 1-2, range: 1-6). As expected, relapse was predicted by use of substances (adjHR: 2.61, 95% CI:1.32-5.15, p=0.0006) and poor adherence to pharmacologic treatment (adjHR 0.38, 95% CI: 0.22-0.65, p<0.0001) (Supplemental table 3).

#### 3.4.4 Rehospitalization

72 patients were hospitalized during follow-up (31.0%). Median time to hospitalization was 266 days (IQR: 90-610) and median number of hospitalizations was 1 (IQR:1-2, range 1-7). Increased rates were associated with use of substances (adjHR:3.39, 95% CI:1.38-8.32, p=0.008), poor adherence to pharmacologic treatment (adjHR:0.35, 95% CI:0.18-0.68, p=0.002), less than 11 years of education (adjHR:0.46, 95% CI:0.26-0.83, p=0.009) and hospitalization at the time of presentation (adjHR:2.52, 95% CI:1.11-5.72, p=0.03) (Supplemental table 4).

## 4. Discussion

This study is one of the few studies conducted in Latin America reporting on longitudinal outcomes of first episode schizophrenia patients. We found that a better response to treatment was associated with shorter DUP, more years of education, less first-order symptoms and being hospitalized at the time of presentation. A shorter DUP and being hospitalized at the time of presentation was also associated with remission of symptoms along with an older age of onset. Predictably, relapse and rehospitalization were both predicted by use of substances and poor adherence to treatment but having less years of education and being hospitalized at the time of presentation also predicted rehospitalization.

There are few points of comparison given the limited number of FEP studies from Latin America, but our finding that remission of symptoms was associated with an older age of onset was similar to the finding by Mena and colleagues (2018) who found that early treatment resistance was associated with younger age of onset. Similarly, our finding that a shorter DUP was associated with better overall response and remission of symptoms is in agreement with the findings by Gonzalez-Valderrama (2017) who found that a shorter DUP was associated with improvement in negative symptoms.

Importantly, the demographic characteristics of the patients in our study are very similar to the demographic characteristics of large RCTs with FEP studies conducted in high income countries. For example, patients included in our study had a median age of 25 y/o, were 73% male, and 90% were living with family and those numbers were comparable to patients in the RAISE study (Kane et al., 2015) (mean age=23 y/o, 66% male, and 71% living with family), and the EUFEST study (Kahn et al., 2008) (mean age=25 y/o, 60% male, and 86% living with family or somebody else). Similarly, when comparing treatment practices between our patients in our study and those included in the RAISE study at the time of enrollment (Robinson et al., 2015), we found that the two most common used antipsychotics in both studies were risperidone and olanzapine, however, the use of olanzapine in our study is much higher compared to the RAISE study (60.2 % vs. 17%), even though olanzapine is not a first line treatment for FEP patients (Kreyenbuhl et al., 2010). Interestingly, the use of clozapine (6.8% vs. 0.7%), mood stabilizers (25.6% vs. 9.2%) and anti-anxiety medications (37.2% vs. 10.4%) was higher in our study compared to RAISE, whereas patients enrolled in RAISE had higher use of antidepressants compared to our study (31.9% vs. 23.9%).

Our findings mirror the findings of high-income countries in regard to predictors of clinical outcomes. We found that a shorter DUP was associated with better response and/or remission and this has been reported by several other studies (Lieberman et al., 1996; Robinson et al., 2004; Perkins et al., 2004; Emsley et al., 2007). Similarly, our finding that a later age of onset was associated with remission was also observed by other investigators (Malla et al., 2006; Crespo-Facorro et al., 2007). Moreover, we replicated the finding that co-morbid substance use and non-adherence to treatment are strongly associated with relapse and rehospitalization (Robinson et al., 1999; Alvarez-Jimenez et al., 2012; Caseiro et al., 2012; Addington et al., 2013). Lastly, our finding that hospitalization at presentation was predictive of rehospitalization was also reported by Addington and colleagues (2013).

Our study has several limitations. Firstly, data was collected as part of routine psychiatric care and therefore it is likely that data was not as carefully collected and entered as if it was a research study. On the other hand, all data was obtained from the EHR, which allowed us to historically review every single visit in every patient. An important limitation is that the definition of our outcomes was based on the text entered by the psychiatrist in the EHR. Even though phrases such as “patient is getting better, patient is improving” are often vague in clinical practice, they do reflect the clinician’s perception of the patient’s clinical condition, which in turn leads to treatment decisions. Strengths of our study include the inclusion of a large first episode schizophrenia sample collected in an urban environment from a lower-middle income (LMIC) country. Furthermore, we had the opportunity to review and obtain longitudinal data from EHR from a large group of patients which allowed us to arrive to clinically relevant outcomes in each individual. Therefore, with this study we demonstrate predictors or clinically relevant outcomes in “real world” conditions in Colombia, South America, in contrast to data obtained from specialized research clinics in academic centers in the US or Europe.

Given that we demonstrated that first episode schizophrenia patients in Medellin, Colombia, have similar demographical and clinical characteristics compared to first episode schizophrenia patients in the US and Europe, research studies could be conducted in this region to either test the feasibility and implementation of interventions developed in high income countries, such as the Navigate intervention, developed as part of the Recovery After Initial Schizophrenia Episode RAISE study (Kane et al., 2015). The successful implementation of these type of interventions in LIMC like Colombia, delivered by a well-trained and highly specialized teams, as is the case in the OnTrackNY program (Nossel et al., 2018), would likely lead to better psychosocial functioning, reduced psychopathology and better quality of life (Nossel et al., 2018).

## Data Availability

All data produced in the present study are available upon reasonable request to the authors

## Acknowledgements

The authors would like to thank Taylor Marzouk for proofreading the manuscript.

## Funding

This research did not receive any specific grant from funding agencies in the public, commercial, or not-for-profit sectors.

## Author contributors

**Garcia, J:** Conceptualization, Methodology, Writing original draft, Formal analysis. **Agudelo, LM**: Conceptualization, Investigation, Writing-Reviewing and Editing. **Canas, MA**: Writing-Reviewing and Editing, Visualization. **Castro-Campos N:** Writing-Reviewing and Editing, Visualization. **Ribero**, O: Investigation, Writing-Reviewing and Editing. **Gallego, J:** Conceptualization, Methodology Writing-Reviewing and Editing, Supervision, Formal analysis.

## Declarations of interest

none

## Notes

### Competing Interest Statement

The authors have declared no competing interest.

### Funding Statement

This study did not receive any funding

### Author Declarations

Ethics committee of Hospital Mental de Antioquia and SAMEIN gave ethical approval for this work

