## Supplemental files (4 tables) for "Predictors of Treatment Response, Remission, Relapse and Rehospitalization in First-Episode Psychosis Patients in Medellin, Colombia"

**Supplemental Table 1. Predictive factors for Response**

| **Factor** | **Response** | **HR crude** | **CI 95%** | **P Value** | **HR Adjusted** | **CI 95%** | **P Value** |
| --- | --- | --- | --- | --- | --- | --- | --- |
|  | Frec. (%) |  |  |  |  |  |  |
| Male vs. Female | 150 (87.2) vs. 51 (85.0) | 1.13 | 0.82 – 1.58 | 0.44 | 1.54 | 0.94 – 2.52 | 0.09 |
| Unemployed vs. employed | 100 (89.3) vs 99 (84.6) | 1.03 | 0.78 – 1.39 | 0.79 | 1.17 | 0.78 – 1.75 | 0.44 |
| Single vs non-single | 162 (86.6) vs 36 (90.0) | 1.11 | 0.76 – 1.64 | 0.58 | 0.78 | 0.47 – 1.28 | 0.33 |
| Education ≥ 11 years vs < 11 years | 96 (88.9) vs 97 (87.4) | 1.39 | 1.04 – 1.87 | 0.03 | 1.51 | 1.01 – 2.24 | *0.04* |
| Middle/ high income vs low income | 122 (89.1) vs 79 (84.0) | 1.24 | 0.92 – 1.66 | 0.16 | 1.06 | 0.71 – 1.57 | 0.78 |
| Urban vs suburban/rural | 190 (87.2) vs 11 (84.6) | 1.03 | 0.55 – 1.96 | 0.92 | 1.25 | 0.45 – 3.48 | 0.67 |
| DUP > 6m vs ≤ 6m | 95 (81.9) vs 97 (93.3) | 0.61 | 0.45 – 0.82 | 0.001 | 0.58 | 0.40– 0.84 | *<0.0001* |
| Age of Onset (Early vs adulthood >18 y) | 53 (84.1) vs 143 (88.3) | 0.94 | 0.68 – 1.31 | 0.75 | 0.94 | 0.65 – 1.36 | 0.74 |
| First-order symptoms vs NP | 128 (84.8) vs 72 (90.0) | 0.71 | 0.52 – 0.96 | 0.03 | 0.52 | 0.34 – 0.81 | *0.004* |
| Negative symptoms vs NP | 165 (86.4) vs 36 (87.8) | 0.79 | 0.54 – 1.15 | 0.22 | 0.76 | 0.47 – 1.24 | 0.27 |
| Disorganized symptoms vs NP | 134 (85.9) vs 67 *88.2) | 0.79 | 0.59 – 1.09 | 0.16 | 0.71 | 0.44 – 1.14 | 0.16 |
| Aggressiveness vs NP | 132 (88.0) vs 69 (84.1) | 1.38 | 1.01 – 1.88 | 0.04 | 0.74 | 0.47 – 1.16 | 0.19 |
| SUD vs NP | 98 (88.3) vs 103 (85.8) | 0.98 | 0.73 – 1.30 | 0.87 | 0.86 | 0.54 – 1.38 | 0.54 |
| Family history of SZ vs NP | 52 (86.7) vs 149 (87.1) | 0.94 | 0.68 – 1.32 | 0.73 | 0.84 | 0.47 – 1.12 | 0.15 |
| Adherence to AP vs non-adherence | 94 (91.3) vs 78 (80.4) | 1.28 | 0,93 – 1.73 | 0.12 | 1.23 | 0.85 – 1.81 | 0.27 |
| Hospitalization at presentation vs none | 141 (91.5) vs 77 (77.9) | 2.48 | 1.77 – 3.49 | < 0.0001 | 3.69 | 2.29 – 5.91 | *<0.0001* |

DUP: Duration of untreated psychosis

NP: Not present

SUD: Substance use disorder

SZ: schizophrenia

AP: Antipsychotic medications

**Supplemental Table 2. Predictive factors for Remission**

| **Factor** | **Response** | **HR crude** | **CI 95%** | **P Value** | **HR Adjusted** | **CI 95%** | **P Value** |
| --- | --- | --- | --- | --- | --- | --- | --- |
|  | Frec. (%) |  |  |  |  |  |  |
| Male vs. Female | 102 (59.3) vs 33 (55.0) | 1.24 | 0.82 – 1.87 | 0.3 | 1.23 | 0.69 – 2.20 | 0.47 |
| Unemployed vs. employed | 63 (56.2) vs 70(59.8) | 1.01 | 0.72 – 1.44 | 0.93 | 0.72 | 0.46 – 1.14 | 0.16 |
| Single vs non-single | 110 (58.8) vs 24 (60.0) | 1.24 | 0.78 – 1.96 | 0.36 | 1.07 | 0.59 – 1.94 | 0.83 |
| Education ≥ 11 years vs < 11 years | 68 (62.9) VS 61 (54.9) | 1.12 | 0.79 – 1.60 | 0.66 | 0.96 | 0.62 – 1.50 | 0.87 |
| Middle/ high income vs low income | 84 (61.3) vs 51 (54.3) | 1.02 | 0.71 – 1.45 | 0.92 | 1.01 | 0.63 – 1.61 | 0.95 |
| Urban vs suburban/rural | 130 (59.6) vs 5 (38.5) | 1.39 | 0.57 – 3.42 | 0.46 | 1.09 | 0.33 – 3.51 | 0.89 |
| DUP > 6m vs ≤ 6m | 55 (47.4) vs 75 (72.1) | 0.48 | 0.34 – 0.69 | <0.0001 | 0.52 | 0.34 – 0.79 | 0.002 |
| Age of Onset (Early vs adulthood >18 y) | 30 (50.8) vs 103 (63.6) | 0.62 | 0.41 – 0.94 | 0.03 | 0.47 | 0.28 – 0.82 | 0.007 |
| First-order symptoms vs NP | 86 (56.9) vs 48 (60.0) | 1.24 | 0.86 – 1.79 | 0.24 | 1.26 | 0.81 – 1.95 | 0.3 |
| Negative symptoms vs NP | 112 (58.6) vs 23 (56.1) | 1.12 | 0.70 – 1.79 | 0.63 | 1.02 | 0.59 – 1.78 | 0.93 |
| Disorganized symptoms vs NP | 89 (57.1) vs 46 (60.5) | 1.26 | 0.87 – 1.83 | 0.21 | 1.09 | 0.67 – 1.77 | 0.73 |
| Aggressiveness vs NP | 84 (56.0) vs 51 (62.2) | 1.07 | 0.74 – 1.53 | 0.72 | 0.69 | 0.42 – 1.13 | 0.14 |
| SUD vs NP | 71 (63.9) vs 64 (53.3) | 1.19 | 0.84 – 1.69 | 0.32 | 1.4 | 0.83 – 2.36 | 0.2 |
| Family history of SZ vs NP | 36 (60.0) vs 99 (57.9) | 0.79 | 0.54 – 1.18 | 0.25 | 0.74 | 0.45 – 1.23 | 0.25 |
| Adherence to AP vs non-adherence | 70 (67.9) vs 57 (58.2) | 1.24 | 0.87 – 1.78 | 0.23 | 1.51 | 0.99 – 2.30 | 0.05 |
| Hospitalization at presentation vs none | 95 (61.3) vs 40 (51.9) | 1.86 | 1.26 – 2.75 | 0.002 | 2.32 | 1.39 – 3.88 | 0.001 |

DUP: Duration of untreated psychosis

NP: Not present

SUD: Substance use disorder

SZ: schizophrenia

AP: Antipsychotic medications

**Supplemental Table 3. Predictive factors for Relapse**

| **Factor** | **Response** | **HR crude** | **CI 95%** | **P Value** | **HR Adjusted** | **CI 95%** | **P Value** |
| --- | --- | --- | --- | --- | --- | --- | --- |
|  | Frec. (%) |  |  |  |  |  |  |
| Male vs. Female | 69 (40.1) vs 19 (31.7) | 1.43 | 0.86 – 2.38 | 0.14 | 1.28 | 0.59 – 2.76 | 0.52 |
| Unemployed vs. employed | 45 (40.2) vs 43 (36.7) | 1.15 | 0.75 – 1.76 | 0.51 | 0.62 | 0.35 – 1.10 | 0.1 |
| Single vs non-single | 75 (40.1) vs 13 (32.5) | 1.48 | 0.82 – 2.67 | 0.19 | 1.18 | 0.57 – 2.45 | 0.65 |
| Education ≥ 11 years vs < 11 years | 43 (39.8) vs 44 (39.6) | 0.88 | 0.57 – 1.34 | 0.54 | 0.89 | 0.54 – 1.49 | 0.68 |
| Middle/ high income vs low income | 56 (40.9) vs 32 (34.0) | 0.92 | 0.59 – 1.43 | 0.71 | 0.93 | 0.56 – 1.58 | 0.81 |
| Urban vs suburban/rural | 86 (39.4) vs 2 (15.4) | 1.98 | 0.49 – 8.08 | 0.34 | 2.29 | 0.49 –10.58 | 0.29 |
| DUP > 6m vs ≤ 6m | 43 (37.1) vs 42 (40.4) | 0.89 | 0.58 – 1.37 | 0.6 | 0.85 | 0.51 – 1.44 | 0.56 |
| Age of Onset (Early vs adulthood >18 y) | 28 (44.4) vs60 (37.0) | 1.19 | 0.75 – 1.89 | 0.75 | 0.9 | 0.52 – 1.57 | 0.72 |
| First-order symptoms vs NP | 53 (35.1) vs 35 (43.7) | 1.06 | 0.69 – 1.65 | 0.77 | 0.89 | 0.49 – 1.61 | 0.7 |
| Negative symptoms vs NP | 73 (38.2) vs 15 (36.6) | 1.2 | 0.68 – 2.09 | 0.52 | 1.15 | 0.60 – 2.19 | 0.67 |
| Disorganized symptoms vs NP | 54 (34.6) vs 34 (44.7) | 0.97 | 0.63 – 1.51 | 0.9 | 0.6 | 0.30 – 1.21 | 0.16 |
| Aggressiveness vs NP | 61 (40.7) vs 27 (32.9) | 1.44 | 0.91 – 2.28 | 0.12 | 1.42 | 0.79 – 2.54 | 0.24 |
| SUD vs NP | 58 (52.2) vs 30 (25.0) | 2.72 | 1.73 – 4.29 | <0.0001 | 2.61 | 1.32 – 5.15 | 0.006 |
| Family history of SZ vs NP | 24 (40.0) vs 64 (37.4) | 0.86 | 0.53 – 1.40 | 0.55 | 1.22 | 0.69 – 2.17 | 0.49 |
| Adherence to AP vs non-adherence | 31 (30.1) vs 56 (57.1) | 0.35 | 0.23 – 0.56 | <0.0001 | 0.38 | 0.22 – 0.65 | <0.0001 |
| Hospitalization at presentation vs none | 68 (43.9) vs 20 (25.9) | 1.9 | 1.12 – 3.20 | 0.02 | 1.16 | 0.62 – 2.16 | 0.64 |

DUP: Duration of untreated psychosis

NP: Not present

SUD: Substance use disorder

SZ: schizophrenia

AP: Antipsychotic medications

**Supplemental Table 4: Predictive Factor for rehospitalization**

| **Factor** | **Response** | **HR crude** | **CI 95%** | **P Value** | **HR Adjusted** | **CI 95%** | **P Value** |
| --- | --- | --- | --- | --- | --- | --- | --- |
|  | Frec. (%) |  |  |  |  |  |  |
| Male vs. Female | 58 (33.7 vs 14 (23.3) | 1.54 | 0.86 – 2.76 | 0.15 | 0.62 | 0.23 – 1.64 | 0.33 |
| Unemployed vs. employed | 41 (36.6) vs 30 (25.7) | 1.59 | 0.99 – 2.57 | 0.05 | 0.55 | 0.29 – 1.03 | 0.06 |
| Single vs non-single | 63 (33.7) vs 8 (20.0) | 1.9 | 0.91 – 3,97 | 0.09 | 1.31 | 0.49 – 3.46 | 0.58 |
| Education ≥ 11 years vs < 11 years | 27 (25.0) vs 42 (37.8) | 0.52 | 0.32 – 0.85 | 0.01 | 0.46 | 0.26 – 0.83 | 0.009 |
| Middle/ high income vs low income | 39 (28.5) vs 32 (34.0) | 0.64 | 0.40 – 1.03 | 0.07 | 0.91 | 0.51 – 1.63 | 0.76 |
| Urban vs suburban/rural | 69 (31.6) vs 2 (15.4) | 1.23 | 0.30 – 5.06 | 0.77 | 0.78 | 0.16– 3.75 | 0.76 |
| DUP > 6m vs ≤ 6m | 31 (26.7) vs 38 (36.5) | 0.71 | 0.44 – 1.15 | 0.24 | 0.74 | 0.42 – 1.31 | 0.29 |
| Age of Onset (Early vs adulthood >18 y) | 24 (38.1) vs 46 (28.4) | 1.35 | 0.82 – 2.23 | 0.23 | 1.19 | 0.66 – 2.17 | 0.56 |
| First-order symptoms vs NP | 51 (33.8) vs 21 (26.2) | 1.59 | 0.96 – 2.66 | 0.07 | 1.55 | 0.74 – 3.24 | 0.24 |
| Negative symptoms vs NP | 61 (31.9) vs 11 (26.8) | 1.22 | 0.64 – 2.33 | 0.54 | 0.69 | 0.31 – 1.52 | 0.36 |
| Disorganized symptoms vs NP | 55 (35.3) vs 17 (22.4) | 2.05 | 1.19 – 3.54 | 0.01 | 0.68 | 0.29 – 1.59 | 0.38 |
| Aggressiveness vs NP | 59 (39.3) vs 13 (15.8) | 3.36 | 1.83 – 6.14 | <0.0001 | 1.92 | 0.94 – 3.91 | 0.07 |
| SUD vs NP | 54 (48.6) vs 17 (14.2) | 3.78 | 2.19 – 6.56 | <0.0001 | 3.39 | 1.38 – 8.32 | 0.008 |
| Family history of SZ vs NP | 13 (20.0) vs 59 (34.5) | 0.44 | 0.23 – 0.83 | 0.01 | 0.63 | 0.30 – 1.30 | 0.21 |
| Adherence to AP vs non-adherence | 17 (16.5) vs 54 (55.1) | 0.27 | 0.15 – 0.46 | <0.0001 | 0.35 | 0.18 – 0.68 | 0.002 |
| Hospitalization at presentation vs none | 64 (41.3) vs 8 (10.4) | 4.9 | 2.35 – 10.24 | <0.0001 | 2.52 | 1.11 – 5.72 | 0.03 |

DUP: Duration of untreated psychosis

NP: Not present

SUD: Substance use disorder

SZ: schizophrenia

AP: Antipsychotic medications
